# Aging and Diabetes: Insights from American Seniors on Self-Care Practices, A Systematic Review & Meta-Synthesis

**DOI:** 10.1101/2024.07.01.24309671

**Authors:** David F. Lo, Ahmed Gawash, Kunal P. Shah, Jasmine Emanuel, Brandon Goodwin, Don D. Shamilov, Gaurav Kumar, Nathan Jean, Christian P. White

**Author notes:** **Email addresses** David F. Lo Ahmed Gawash Kunal P. Shah, Jasmine Emanuel Brandon Goodwin Don D. Shamilov Gaurav Kumar, Nathan Jean Christian P. White. **Corresponding author:** David F. Lo, MBS Department of Medicine, Rowan School of Osteopathic Medicine Address: 1 Medical Center Dr, Stratford, NJ 08084. **Funding:** No funding was received for this study/paper. **Competing interests:** The authors declare that they have no competing interests.

## Abstract

This review aims to unravel the intricacies of diabetic self-management among geriatric people, drawing on current insights and understanding the complex paths geriatric people navigate. A wide search was conducted in health-oriented databases, including CINAHL, Embase, PsycINFO, MEDLINE, PubMed, Web of Science, and Cochrane Library while gray literature was excluded. The search combined keywords and subject headings, focusing on the geriatric population, diabetes, self-management, and qualitative research. A three-tiered screening process was employed, with titles and then abstracts initially reviewed. Full-text analysis followed, with disagreements resolved among reviewers. In total, there were 248 participants included across these eight studies. Positive attitudes and perceptions were found to play a significant role in optimizing diabetes self-care outcomes. Support from family and friends was identified as crucial for self-care, while healthcare professionals often lacked adequate support and encouragement. Participants emphasized the importance of listening to their bodies and acknowledging hidden issues. These themes collectively highlight the multifaceted aspects of diabetes self-care and the impact of various factors on the self-management experiences of geriatric individuals with diabetes. The goal of this review is not to objectify self-management as a treatment strategy but to emphasize the importance of cultivating positive attitudes, respecting individual values, and addressing cultural and ethnic differences in healthcare practices to enhance self-management in this population. By embracing cultural diversity, understanding barriers, and respecting individual values, healthcare professionals and policymakers can improve the quality of life for the geriatric population living with diabetes.

## Introduction

Understanding the management of Diabetes Mellitus (DM) is of paramount importance, given its status as the 8th leading cause of death in the United States (U.S.) population and the primary cause of kidney failure, lower-limb amputation, and adult blindness (1). Despite advancements in medical knowledge and innovative treatment outcomes, the associated costs pose challenges for many individuals, particularly the geriatric population, who may lack the necessary financial or emotional support (17). Estimates project a significant rise in the prevalence of DM among the U.S. population, with numbers expected to increase from 22.3 million in 2014 to 39.7 million in 2030 and a staggering 60.6 million in 2060 (2). This growing aging population in the United States is increasing the challenge of managing Type 2 diabetes (T2D). By 2060, an estimated 95 million people 65 and older will be affected by T2D (3). Additionally, the US demographic landscape is shifting, with a significant increase in diversity from 2010 to 2020.

Over the past decade, the population diversity index has increased from 54.9% to 61.1% (43). This change reflects ongoing trends in population dynamics, indicating a continued rise. According to the US Census Bureau, this shift is driven by the aging baby boomer generation and increased immigration, both of which contribute to population growth and diversification (44, 45). This prompts the question of how to best prepare and assist this population in effectively managing chronic conditions. Recent strides in medical advancements, technological breakthroughs, and healthcare delivery have led to a remarkable extension in human lifespans. This has resulted in a sharp surge in the global population of seniors (4), with an expected one in six people in the world over age 65 by 2050 (5). The global trend of aging populations has placed a significant and challenging burden on healthcare systems worldwide (6). Throughout the paper, varying terms have been utilized to describe older adults (i.e., “elderly people,” “geriatric,” “older individuals,” and “elderly individuals”). This demonstrates a nuanced understanding of language’s impact on inclusivity and precision as well as catering to diverse audiences/contexts while fostering respect for individual preferences. It also reflects the need for a broader search among papers when performing the meta-synthesis.

As people live longer and adopt more urban lifestyles, T2D has become more common among geriatric people (7). Unfortunately, there’s no cure for diabetes, and it’s a major health challenge of our time (1). It leads to various complications, including diabetic ketoacidosis, low blood sugar, heart problems, eye issues, kidney problems, circulatory troubles, foot conditions, and complications during pregnancy (8). The American Diabetes Association (ADA) noted that geriatric people with diabetes face a higher risk of losing their independence, early death, muscle loss, heart diseases, strokes, and high blood pressure compared to those without diabetes (9). Diabetes has a more significant impact on older adults, leading to more disability and higher death rates (10). These challenges place a substantial burden on social services, healthcare systems, and economies (11).

Poor diabetes management can lead to serious and lasting complications, impacting various bodily systems. These complications include vascular disorders, neuropathies, disabilities, diabetes-related distress, depression, and even death (9). Hence, maintaining effective diabetes self-management and enhancing socio-psychological functions are critical to preventing the aforementioned severe outcomes (11). This involves not only medical care but also personal care behaviors such as maintaining a healthy diet, engaging in suitable physical activity, monitoring blood glucose levels, adhering to prescribed medication regimens, and conducting self-foot checks - all important components of diabetes management. (12).

Even with comprehensive measures in place, structured education presents significant challenges. These issues are especially important considering potential demographic shifts as estimated by the Population Reference Bureau (3). Previous research has indicated that self-efficacy, knowledge, social support, self-regulation, and outcome expectations play pivotal roles in influencing self-management among geriatric people with diabetes (5,14). This population is more susceptible to issues like polypharmacy, falls, cognitive impairment, depression, and incontinence compared to those without diabetes, further complicating their care (9,15). These factors underscore the importance of self-management while reinforcing the need for increased support for the geriatric population, whether it be through their healthcare providers or family members tasked with delivering high-quality care to the growing population of geriatric diabetic patients (16,17). The intricacy of self-management thus becomes a crucial and intricate aspect of the lives of the geriatric population coping with diabetes.

Understanding the experience of older adults with T2D is crucial because of other chronic comorbidities present among much of the population (18). These older adults require specialized care. This type of qualitative study helps to create tailored care plans that address their specific needs, enhance communication between healthcare providers, patients, and their families, improve care practices, influence policy and program development, and inspire further research to improve health outcomes. While studies have examined diabetes self-management (such as Chester et al. 2018), there has been limited focus on the unique experience of the older population. Although some research has explored this area, our study specifically is focused on the U.S. due to its distinct cultural, socioeconomic, and healthcare differences (26,27,28,29,30,21,32,42). This review aims to uncover the complexities of self-management among older adults with diabetes in the U.S., drawing on current research and insights.

While several countries, including Ethiopia (20) and Nigeria (21), have delved into the subject of self-management of diabetes, none have explored the precise themes and challenges addressed in our study such as the impact of cultural diversity on self-management, the role of family and social support, the psychological attitudes towards diabetes, and the barriers faced by older adults with T2D in a aging U.S. population. While a review published in 2022 (40) explored the experiences of the elderly with diabetes care, our review distinguishes itself by introducing novel themes, focusing exclusively on subjects in America, and providing an updated perspective specifically on T2D. This paper aims to provide a systematic review and meta-synthesis, a qualitative type of meta-analysis that contributes insights and perspectives previously underrepresented in the existing literature. Qualitative research methodologies are uniquely suited to delve into the subjective perspectives, meanings, and nuances of individuals’ lived experiences. Unlike quantitative approaches, which often prioritize measurable outcomes and generalizability, qualitative methods allow us to explore the complexities, emotions, and contextual factors that shape how individuals perceive and navigate their realities.

In the context of self-management and related areas, such as healthcare interventions or chronic illness management, understanding the lived experience is important. Many aspects of self-management, such as coping strategies, decision-making processes, and adaptation to illness, are deeply personal and influenced by a myriad of factors beyond simply the effectiveness of interventions or the presence of certain symptoms. By focusing on qualitative studies, this review acknowledges the importance of capturing the rich, nuanced narratives of individuals directly impacted by self-management practices or interventions. Bringing all this information together is critical, treating T2D just with a pharmacological approach is not enough, and self-care/management plays an equally important role in managing diabetes health reasons mentioned throughout the paper. These narratives offer invaluable insights into the subjective realities of those managing chronic conditions or navigating healthcare systems, shedding light on their unique challenges, successes, barriers, and facilitators. This study offers an insightful understanding of the challenges and opportunities surrounding geriatric self-management of diabetes as well as the quality of current research.

## Methods

The following systematic review strictly adhered to the PRISMA guidelines, but due to low study numbers and the nature of qualitative studies, a meta-synthesis was conducted instead of a meta-analysis (22). This investigation adhered to the guidelines stipulated by Enhancing Transparency in Reporting the Synthesis of Qualitative Research (ENTREQ) (23). The Critical Appraisal Skills Program (CASP) (24,25) served as the benchmark for the quality assessment of the selected publications.

**Fig 1.**
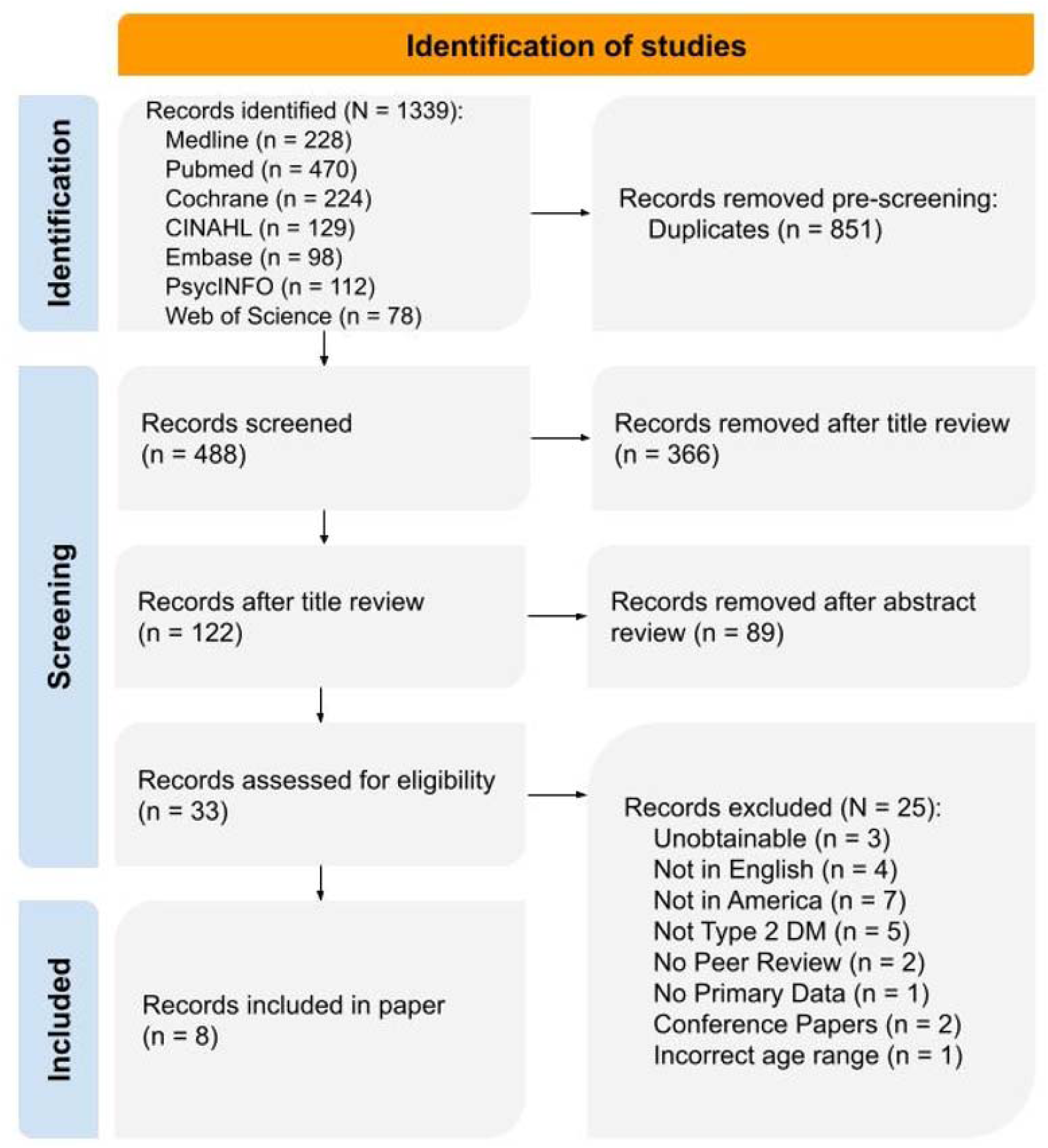
PRISMA flowchart illustrating the systematic review process for Self-Management of Type 2 Diabetes in the Geriatric population in America. The flowchart outlines the selection and screening of studies, including search strategies, eligibility criteria, and the final inclusion of relevant studies for data synthesis and analysis.

### Search Strategy

Primary citation exploration involved several health-oriented databases, including CINAHL, Embase, PsycINFO, MEDLINE, PubMed, Web of Science, and Cochrane Library. All electronic database searches were conducted on January 26, 2024. In addition to electronic queries, the search process included a review of references and manual searches to identify relevant research findings. MeSH terms were used, focusing on key concepts such as Type 2 diabetes, elderly, self-care, qualitative methodologies, and America. The full search string, along with boolean operators, is included below:

(“Type 2” OR “T2” OR “Non-Insulin-Dependent” OR “Adult-Onset”) AND (“Diabetes Mellitus” OR “DM” OR “Diabetes” OR “Diabetic” OR “Hyperglycemia” OR “Glucose Disorder” OR “Metabolic Disorder”) AND (“Aged” OR “Elderly” OR “Geriatric” OR “Older” OR “Old Age” OR “Senior” OR “Elderly”) AND (“Personal Care” OR “Self-Care” OR “Self-Support” OR “Self-Management” OR “Self-Monitoring” OR “Self-Supervision”) AND (“Qualitative” OR “Descriptive” OR “Interpretive” OR “Subjective”) AND (“Interview” OR “Research” OR “Study” OR” Narrative” OR “Experience”) AND (“America” OR “US” OR “USA” OR “United States” OR “United States of America”).

### Inclusion Criteria

The inclusion criteria for paper selection encompassed qualitative studies exploring the experiences, needs, perspectives, and attitudes of geriatric people affected by T2D in relation to the self-management of their condition. Participants included were diagnosed with T2D diabetes, aged 60 and above. Only studies conducted in America were included, as other countries have different cultures, socioeconomic factors, and healthcare systems. Studies between 2009 and 2024 were included. Although some papers are older than 10 years, our study focused on underlying factors and beliefs that still hold true today as they did in the past.

### Exclusion Criteria

Studies not in English were excluded to ensure a consistent language for analysis. Furthermore, we excluded studies that didn’t pertain to T2D since our review aimed to provide insights specifically related to this diabetes subtype. To maintain the integrity of the review, studies without peer review were also excluded, as peer-reviewed articles are generally subject to a rigorous evaluation process that ensures their scientific validity and quality. Gray literature was excluded as well.

We also excluded studies that did not provide primary data, as our objective was to assess the primary research conducted in this field. Secondary evidence, such as reviews, was omitted from our review to maintain a focus on original research and firsthand accounts of experiences and perspectives related to diabetes self-management. Next, studies in which participants were under 60 years old were excluded. Finally, conference papers were excluded from our analysis to ensure that the studies included in the review met established publication standards and underwent a peer-review process. Furthermore, we excluded studies that were not conducted in the U.S. due to unique diet, cultural, and sociological factors. Studies older than 15 years old were excluded due to recent advancements in medical care.

### Selection Process

The initial database query yielded 1,339 results, of which 851 were duplicates. The remaining articles underwent a three-phase screening process where papers were reviewed based on titles and abstracts, excluding 455 more articles. Any disagreements were discussed and resolved between the two screeners (D.F.L. and A.G.); those that weren’t were subjected to a full-text analysis. This analysis resulted in the exclusion of 25 more papers. Any remaining discrepancies were discussed in-depth, and if necessary, a third reviewer would step in to resolve them, but this was not needed.

### Quality Appraisal

To gauge the confidence level of the findings, the confidence ratings were set as high, moderate, or low. The CASP framework was used to assess any methodological limitations among the studies included (24,25). The researchers (D.F.L. and A.G.) evaluated each article independently, and then discussed and compared their assessments to reach a shared conclusion, thus enhancing the reliability of their evaluations.

### Synthesis and analysis

In this stage, the data from the selected studies was subject to in-depth analysis. This model facilitated a large examination of the data, allowing for the extraction of key themes and insights. Table 1 presents a systematic evaluation of several research studies using the Critical Appraisal Skills Program (CASP) scoring system. Some achieved "High" quality status (27,31,32), while others fell into the "Medium" (26,29,30) categories. This minor variability in quality implies that there is occasional inconsistency in the rigor and thoroughness of research in the field.

**Table 1.** Critical appraisal skills program (CASP) score.

|  | Washington & Wang-Letzkus (2009)<sup>26</sup> | Chlebowy et al. (2010)<sup>27</sup> | George & Thomas (2010)<sup>28</sup> | Kirk et al. (2011)<sup>29</sup> | Beverly et al. (2014)<sup>30</sup> | Joo & Lee (2016)<sup>31</sup> | Bustillos & Sharkey (2020)<sup>32</sup> | Narindrarangura et al. (2022)<sup>42</sup> |
| --- | --- | --- | --- | --- | --- | --- | --- | --- |
| Clear research aims? | 1 | 1 | 1 | 1 | 1 | 1 | 1 | 1 |
| Appropriate methodology? | 1 | 1 | 1 | 1 | 1 | 1 | 1 | 1 |
| Design aligned with aims? | 1 | 1 | 1 | 1 | 1 | 1 | 1 | 1 |
| Suitable recruitment? | 0.5 | 1 | 1 | 1 | 0 | 1 | 1 | 1 |
| Relevant data collection? | 1 | 1 | 1 | 1 | 1 | 1 | 1 | 1 |
| All party relationships considered? | 0 | 1 | 1 | 0.5 | 0 | 0 | 1 | 1 |
| Ethical issues considered? | 0 | 1 | 1 | 0 | 0 | 1 | 0 | 1 |
| Rigorous data analysis? | 0 | 1 | 1 | 0.5 | 1 | 1 | 1 | 1 |
| Clear findings? | 1 | 1 | 1 | 1 | 1 | 1 | 1 | 1 |
| Research value assessed? | 1 | 1 | 0.5 | 0.5 | 1 | 0 | 1 | 1 |
| **Score** | **6.5** | **10** | **9.5** | **7.5** | **7** | **8** | **9** | **10** |
| **Quality** | **Med** | **High** | **High** | **Med** | **Med** | **High** | **High** | **High** |

GRADE (1: Yes; 0: No; 0.5: Cannot tell); QUALITY (Low: [0-2.5], Med: [3-7.5], High: [8-10])

Certain aspects exhibit a degree of consistency across the studies. For instance, all studies appear to have clear research aims and appropriate methodologies, receiving a score of 1 in these categories. Certain aspects exhibit a degree of consistency across the studies. For instance, all studies appear to have clear research aims and appropriate methodologies, receiving a score of 1 in these categories. Additionally, all the study designs aligned well with the proposed research objectives. This shows that researchers are careful to ensure their research is well structured. However, ethical considerations vary significantly among these studies. Some align very well (27,28,31), while others don’t even mention any ethical issues (26,29,30,32). This emphasizes how important it is to prioritize ethics in research to protect participants and maintain credibility.

## Results

### Study Selection

Eight qualitative studies were identified and met the inclusion criteria. All of these studies were conducted in the United States and were published in English. While these studies had varying stated focuses and aims, they all shared a common theme of exploring the experiences of geriatric people with diabetes concerning self-management. In total, there were 248 participants included across these eight studies, with participant numbers ranging from 10 to 31 in most studies. The target population for all these studies was elderly, with an age range of 60 to 85 years. All participants had been diagnosed with T2D. For data collection, three studies utilized focus group interviews, three conducted individual interviews, and one employed a combination of both focus group and individual interviews. These studies used a variety of qualitative research methods, including thematic analysis in three studies, phenomenology analysis in one, and content-based analysis in five. A summary of the included studies is provided in Table 2.

**Table 2.** Summary of Studies.

| Author | Year | Objective |
| --- | --- | --- |
| Washington & Wang-Letzkus<sup>26</sup> | 2009 | Identify risk factors related to lifestyle, attitudes, and health beliefs, and the influence on self-care practices of Chinese-Americans |
| Chlebowy et al.<sup>27</sup> | 2010 | Explore self-management practices and challenges in geriatric African American T2D patients. |
| George & Thomas<sup>28</sup> | 2010 | Elucidate experiences and perceptions of individuals with diabetes with regard to self-management, as narrated by older people diagnosed with insulin-dependent diabetes living in a rural area |
| Kirk et al.<sup>29</sup> | 2011 | Examine the emotional and physical symptoms experienced by older diabetes patients in relation to high and low blood sugar levels. |
| Beverly et al.<sup>30</sup> | 2014 | Investigate the values and preferences of older adults with regard to T2D attention |
| Joo & Lee<sup>31</sup> | 2016 | Look for barriers of self-management of diabetes among first-generation Korean-American geriatric immigrants living with T2D in midwest America |
| Bustillos & Sharkey<sup>32</sup> | 2020 | Study various challenges of T2D self-management among older adults who are unable to leave their homes and who receive home-delivered meals regularly |
| Narindrarangura et al.<sup>42</sup> | 2022 | Determine information needed for diabetic patients by utilizing surveys and focus groups to develop improved patient care |

The listed studies cover a wide range of research objectives in the context of diabetes management and self-care among diverse populations. In 2009, Washington and Wang-Letzkus focused on Chinese-American immigrants, aiming to identify risk factors related to lifestyle, attitudes, and health beliefs, and their impact on self-care practices. In 2010, Chlebowy et al. along with George and Thomas explored self-management practices and delved into the perceptions of geriatric people with insulin-dependent diabetes, respectively, shedding light on their self-management narratives. The next year, Kirk et al. 2011, examined the emotional and physical symptoms experienced by older diabetes patients in relation to high and low blood sugar levels. Beverly et al. conducted research in 2014 to investigate the values and preferences of older adults concerning T2D care. In 2016, Joo and Lee studied barriers to self-management among Korean-American geriatric immigrants with T2D in the Midwest. In 2020, Bustillos and Sharkey examined the challenges faced by older adults who receive home-delivered meals and have limited mobility in managing T2D. Finally, in 2022, Narindrarangkura et al. found that only 60% of providers who participated in their study utilized diabetes self-management and support guidelines (DSMES). These studies collectively contribute to a better understanding of the multifaceted aspects of diabetes self-care within specific populations and circumstances, as shown in Table 3.

**Table 3.** Study Characteristics.

| Author | Sample size<br>(N = 248) | Age | Ethnicity | Method |
| --- | --- | --- | --- | --- |
| Washington & Wang-Letzkus (2009)<sup>26</sup> | 13 (6 Females) | >65 | Chinese-American | Focus group interviews |
| Chlebowy et al. (2010)<sup>27</sup> | 38 (27 Females) | >65 | African-American | Focus group interviews |
| George & Thomas (2010)<sup>28</sup> | 10 | 65-85 | White-predominant | Unstructured 1-on-1 interviews |
| Kirk et al. (2011)<sup>29</sup> | 75 (39 Females) | >65 | White-predominant | Structured 1-on-1 interviews |
| Beverly et al. (2014)<sup>30</sup> | 25 | >60 | White-predominant | Focus group interviews |
| Joo & Lee (2016)<sup>31</sup> | 23 (11 Females) | >65 | Korean-American | Focus group interviews & 1-on-1 interviews |
| Bustillos & Sharkey (2020)<sup>32</sup> | 31 (23 Females) | >65 | White-predominant | Semi-structured 1-on-1 interviews |
| Narindrarangku<br>ra et al. (2022)<sup>42</sup> | 33 (18<br>females) | 65-82 | White-predominant | Survey & focus group<br>interviews |

In terms of study characteristics, Washington & Wang-Letzkus’ research involved 13 Chinese-American participants, predominantly over the age of 65, and employed focus group interviews, while Chlebowy focused primarily on African-American patients. George & Thomas conducted a study with 10 participants, primarily from a white-predominant population, aged 65-85, utilizing unstructured interviews. Kirk et al. conducted structured 1-on-1 interviews with 75 participants from a predominantly white demographic group, all aged over 60. In contrast, Beverly et al. examined 25 participants from a similar demographic but used focus groups as their primary research approach. Joo & Lee’s research focused on 23 Korean-American participants, most of whom were over 65 years old. They gathered data through a combination of focus groups and one-on-one interviews. Bustillos & Sharkey’s research involved 31 participants, primarily from a white-predominant population, aged over 65, with data collected through semi-structured interviews. Lastly, Narindrarangkura et al. included 33 participants who were all non-Hispanic and composed of focus groups and survey participants. These variations in sample sizes, participant demographics, and research methods collectively contribute to an insightful understanding of diabetes self-management within diverse populations.

Table 4 summarizes the analysis types and key outcomes of various studies. In Washington & Wang-Letzkus’ 2009 study, a thematic analysis revealed that positive perceptions and optimistic attitudes play a pivotal role in optimizing diabetes self-care outcomes, while Chlebowy et al. 2010 adopted a content-based analysis looking at support from family and friends for diabetes self-care. George & Thomas’ 2010 study employed a phenomenological approach, uncovering the profound experiences and thoughts of individuals with diabetes, from "Your body will let you know" to "The only way out is to die." While Kirk et al. 2011, used content-based analysis to differentiate between symptoms of high and low blood sugar, Beverly et al.’s study used thematic analysis, emphasizing the importance of respecting individual values and preferences in the physician-patient relationship to foster collaboration and confidence in older adults. Joo & Lee’s 2016 study conducted a content-based analysis, identifying various factors impacting diabetes self-management, including the high cost of care, language barriers, and limited access to resources. Bustillos & Sharkey’s 2020 study, employing thematic analysis, delved into the perceived seriousness of diabetes, self-management, and barriers such as physical activity and economic concerns. Finally, Narindrarangkura’s 2022 study, which utilized a content-based approach, indicated that the readability of notes is the main barrier for patients.

**Table 4.**
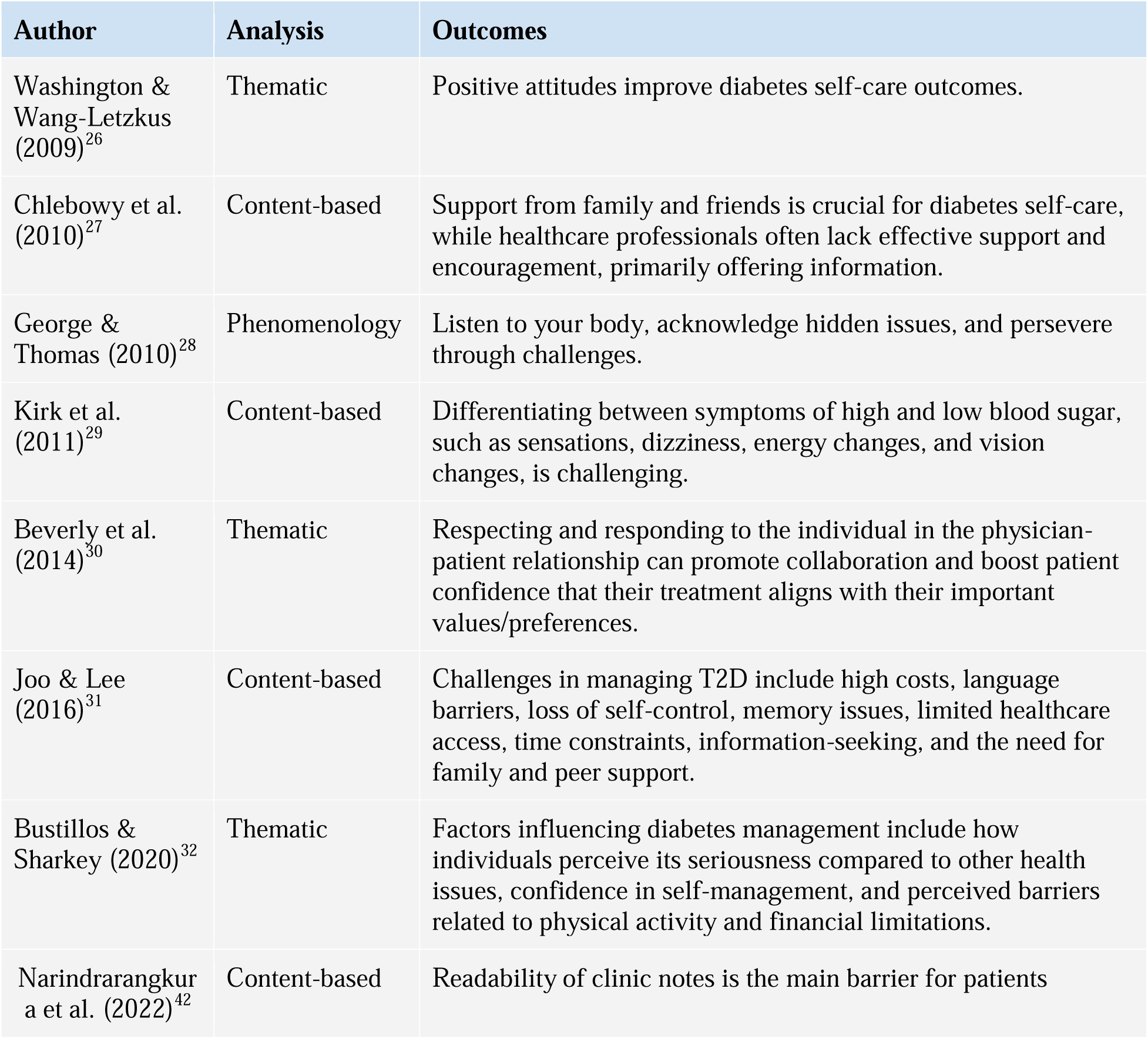
Study Outcomes.

| Author | Analysis | Outcomes |
| --- | --- | --- |
| Washington & Wang-Letzkus (2009)<sup>26</sup> | Thematic | Positive attitudes improve diabetes self-care outcomes. |
| Chlebowy et al. (2010)<sup>27</sup> | Content-based | Support from family and friends is crucial for diabetes self-care, while healthcare professionals often lack effective support and encouragement, primarily offering information. |
| George & Thomas (2010)<sup>28</sup> | Phenomenology | Listen to your body, acknowledge hidden issues, and persevere through challenges. |
| Kirk et al. (2011)<sup>29</sup> | Content-based | Differentiating between symptoms of high and low blood sugar, such as sensations, dizziness, energy changes, and vision changes, is challenging. |
| Beverly et al. (2014)<sup>30</sup> | Thematic | Respecting and responding to the individual in the physician-patient relationship can promote collaboration and boost patient confidence that their treatment aligns with their important values/preferences. |
| Joo & Lee (2016)<sup>31</sup> | Content-based | Challenges in managing T2D include high costs, language barriers, loss of self-control, memory issues, limited healthcare access, time constraints, information-seeking, and the need for family and peer support. |
| Bustillos & Sharkey (2020)<sup>32</sup> | Thematic | Factors influencing diabetes management include how individuals perceive its seriousness compared to other health issues, confidence in self-management, and perceived barriers related to physical activity and financial limitations. |
| Narindrarangkur et al. (2022)<sup>42</sup> | Content-based | Readability of clinic notes is the main barrier for patients |

In Table 5, we provide an overview of the key themes that emerged from the reviewed studies. These themes shed light on various aspects of geriatric self-management of T2D. "Body Signal Awareness" emphasizes the importance of being in tune with one’s body signals, which is fundamental to health and well-being. "Diabetes Care Knowledge & Understanding" highlights the significance of comprehensive knowledge about diabetes care, including medication management, blood glucose monitoring, and preventive measures. "Motivation & Support Systems" underscores the need for robust support systems, encompassing emotional, social, and healthcare support. "Functional Decline Management" addresses the challenges and strategies for managing functional decline, especially in geriatric people. "Psychosocial Attitudes Toward Diabetes" delves into the psychological and emotional aspects of living with diabetes. Finally, "Challenges in Lifestyle" explores the difficulties and obstacles individuals face in adopting and maintaining a healthy lifestyle, particularly in the context of diabetes. These themes collectively contribute to a nuanced understanding of geriatric self-management of T2D.

**Table 5.** Overview of themes.

| Themes | Objectives | No. of studies |
| --- | --- | --- |
| Body Signal Awareness | Importance of being attuned to and responsive to one's body signals is a fundamental aspect of health and well-being. | 3 |
| Diabetes Care Knowledge & Understanding | Importance of acquiring comprehensive knowledge about diabetes care, including medication management, blood glucose monitoring, and preventive measures. | 5 |
| Motivation & Support Systems | Highlighting the need for a robust support system, this theme emphasizes the significance of emotional, social, and healthcare support for individuals living with diabetes. | 3 |
| Functional Decline Management | Addresses the challenges and strategies for managing functional decline that may be associated with diabetes, especially in geriatric people. | 5 |
| Psychosocial Attitudes Toward Diabetes | Explores the psychological and emotional aspects of living with diabetes, including stigma, self-perception, and the impact of attitudes on self-care. | 4 |
| Challenges in Lifestyle | Explores the difficulties and obstacles individuals face when trying to adopt and maintain a healthy lifestyle, particularly in the context of diabetes. | 3 |

To effectively manage diabetes, a multifaceted approach addressing psychological, social, and healthcare needs is essential. Personalized psychological support is crucial for addressing anxiety, depression, and emotional adaptation to diabetes, utilizing cognitive-behavioral techniques to enhance stress management and coping skills. Regular psychological assessments should be integrated into healthcare appointments to monitor and support emotional well-being. Providing educational materials and training in coping skills further aids in managing the emotional aspects of diabetes. Social support is equally important, with peer support groups and culturally competent education for providers helping to alleviate feelings of isolation. Including sociocultural elements such as religious values in the assessment process and directing individuals to relevant support services can enhance social integration (40). Establishing a compassionate rapport with patients, delivering clear and comprehensive information about their condition, and addressing individual anxieties can significantly improve emotional well-being in healthcare settings. Tailoring reassurance to address specific concerns and providing pertinent information and support are key components of effective diabetes management (46). Together, the social, mental, and healthcare needs, along with the specific clinical actions and recommendations, are summarized in Table 6.

**Table 6.** Social, Mental, and Healthcare Needs for Diabetes Self-management in the Elderly.

|  | Specific actions | Clinical recommendations |
| --- | --- | --- |
| **Psychological** | Individualized support<br>Cognitive-behavioral strategies<br>Regular psychological check-ins<br>Education and coping skills | Provide personalized psychological assistance to address anxiety, depression, and emotional adaptation to diabetes. Utilize cognitive-behavioral techniques for stress management, coping, and emotional fortitude. Integrate regular psychological assessments into healthcare appointments to observe and attend to emotional well-being. Supply educational materials and training in coping skills for the management of the emotional dimensions of diabetes (40). |
| **Social** | Provide peer support groups and educate providers on cultural competence | Offer normalization and peer support groups to alleviate feelings of isolation. Include sociocultural elements like religious values and beliefs in the assessment process. Direct individuals to relevant support services when needed (40). |
| **Healthcare** | Reassurance and containment | Establish a robust and compassionate rapport with the patient, expressing authentic care and empathy. Deliver precise and comprehensive information regarding the patient's condition, treatment, and healthcare procedures. Acknowledge that every patient might harbor distinct |
|  |  | anxieties and apprehensions. Tailoring reassurance to cater to their specific concerns and furnishing pertinent information and support can notably improve their emotional well-being (46). |

## Discussion

This systematic review and meta-synthesis presents an insightful understanding of geriatric self-management of T2D in America. In line with existing research, our findings underscore the critical role of optimism and self-efficacy in improving self-management outcomes among the elderly. This aligns with previous studies that have highlighted the importance of a positive attitude in dealing with chronic illnesses (34). This discussion delves into the unique perspectives on geriatric self-management of T2D, emphasizing cultural diversity, barriers, positive attitudes, and the crucial role of the physician-patient relationship. It provides practical implications for healthcare practice and offers recommendations for healthcare professionals to enhance geriatric diabetes management. The limitations and the need for future research are acknowledged, with clinical implications underscoring the importance of a positive and empowering environment, cultural sensitivity, and tailored support for geriatric patients (40).

### Unique Perspectives on Geriatric Self-Management

Geriatric self-management of T2D stands apart due to the unique confluence of factors faced by geriatric people. Cultural diversity among the elderly, as demonstrated in the reviewed studies, plays a pivotal role in shaping self-management strategies. Geriatric individuals, who may be rooted in traditions, have different perceptions of their condition and its management (40).

### Barriers to Self-Management

The studies have revealed several barriers to T2D self-management faced by geriatric individuals. High costs, language barriers, limited access to healthcare, and memory issues (31,32) are among the challenges that need to be addressed. Moreover, time constraints and the need for family and peer support play significant roles in shaping their self-management experiences (27,31). Understanding these barriers is essential for healthcare professionals and policymakers to develop targeted interventions.

### Positive Attitudes and Strategies

Optimism and self-efficacy are not only powerful assets for geriatric self-management (26) but also critical components of a holistic healthcare approach. Cultivating positive attitudes among geriatric individuals can lead to better adherence to treatment plans and self-care behaviors. Strategies to enhance self-efficacy should be central to interventions designed to improve the self-management of T2D in the geriatric population.

### Physician-Patient Relationship

The importance of respecting individual values and preferences in the physician-patient relationship cannot be overstated. The reviewed studies indicate that fostering collaboration and trust in older adults by aligning treatment with their values and preferences can be a catalyst for improved self-management (30). Healthcare providers should prioritize effective communication and shared decision-making to facilitate this vital aspect of care (27).

### Cultural and Ethnic Differences

The cultural and ethnic diversity highlighted in the reviewed studies (30) calls for a more nuanced approach to geriatric diabetes management. To ensure healthcare professionals are equipped to deliver culturally sensitive care, cultural competency training is essential. This training should encompass cultural sensitivity, language proficiency, and a deeper understanding of the specific healthcare challenges faced by different cultural groups. Healthcare providers should undergo such training to enhance their ability to provide care that respects the cultural backgrounds and beliefs of geriatric patients. These perspectives must be integrated into healthcare practices to create culturally sensitive interventions (30).

### Implications for Healthcare Practice

This systematic review offers practical implications for healthcare professionals, policymakers, and practitioners working with geriatric patients with T2D. Comprehensive care models should encompass culturally sensitive education (30), improved access to affordable care (32), and proactive memory support (31). Encouraging family and peer involvement in the care process can further bolster self-management outcomes (31).

### Recommendations for Healthcare Professionals

1. Cultural Competency Training: Healthcare professionals should undergo cultural competency training to better understand and respect the cultural backgrounds and beliefs of geriatric patients. This training should encompass cultural sensitivity, language proficiency, and a deeper understanding of the specific healthcare challenges faced by different cultural groups.
2. Address Barriers: Healthcare professionals should proactively address barriers to self-management, including high costs, language barriers, limited access to healthcare, and memory issues. This may involve providing language support, improving access to affordable care, and simplifying medication regimens.
3. Promote Positive Attitudes: Cultivating positive attitudes among geriatric individuals is crucial. Healthcare professionals should share success stories, offer continuous encouragement, and promote hope to enhance geriatric patients’ commitment to self-management.
4. Respect Individual Values: The physician-patient relationship should prioritize respecting individual values and preferences. Tailoring treatment plans to align with each patient’s values is essential for fostering collaboration and trust.
5. Tailored Support: Recognize the unique emotional and practical needs of geriatric individuals with diabetes. Tailored support groups should be designed to address these needs, including the psychological and social dimensions of diabetes.

### Limitations and Future Research

While our review provides valuable insights into geriatric self-management of T2D, it is essential to acknowledge its limitations and, importantly, explain why they may not be as restrictive as they initially appear. Few of the included studies predominantly focused on specific ethnic and cultural groups; however, experiences of self-care among older patients with diabetes may be shared among cultures to some extent. We recognize the need for more diverse studies that encompass a wider range of ethnic and cultural contexts to provide a more in-depth understanding of geriatric self-management of diabetes. Furthermore, we did not delve deeply into the impact of cognitive awareness and memory on self-management in geriatric patients, which can be seen as a limitation. However, this particular aspect of self-management is an area that requires more focused attention and research. Future studies should explore how cognitive factors may influence the self-care practices of geriatric individuals with T2D to better inform tailored interventions and support mechanisms.

Furthermore, the rigor of data analysis and the assessment of research value exhibit disparities among the studies. While some studies meticulously analyze data and offer valuable contributions (27,32), others only partially meet these criteria (29,30). This necessitates for researchers to prioritize rigorous data analysis and clear communication of findings to enhance the credibility and significance of their research.

### Clinical Implications

The findings of this systematic review and meta-synthesis offer valuable insights that have significant clinical implications for healthcare practitioners, patient care, and healthcare policy, especially in the context of geriatric self-management of T2D. Cultivating a positive and empowering environment is crucial in enhancing self-management outcomes among geriatric patients. Healthcare professionals play an instrumental role in creating a patient-centered care atmosphere, fostering optimism, and empowering patients with a sense of self-efficacy. Sharing success stories, offering continuous encouragement, and promoting hope are straightforward yet potent strategies that can aid in bolstering geriatric patients’ commitment to self-management (26). Recognizing and respecting the cultural diversity among geriatric patients is another imperative clinical implication (30). Healthcare practitioners should take into account each patient’s cultural background, values, and individual preferences when designing care plans (30). This knowledge should be the foundation for tailoring care to meet the unique needs of each patient. Addressing language barriers through interpreter services and multilingual materials is also essential to ensure equitable access to information and healthcare services for all patients, irrespective of their linguistic background.

Memory challenges, which are not uncommon among geriatric individuals, must be effectively addressed in clinical practice. Clinicians should incorporate memory-friendly strategies into diabetes management plans. Simplifying medication regimens, providing easy-to-understand written instructions, and employing reminders through phone applications or the support of caregivers can significantly enhance medication adherence and self-care practices (35). Moreover, the economic barriers associated with managing T2D should be tackled at the healthcare policy level. High costs can pose a substantial barrier to geriatric patients, so policies should explore opportunities to alleviate the financial burden. This may include increasing access to affordable medications, improving insurance coverage, and providing financial support for necessary supplies and equipment (36).

Family and peer support should not be underestimated. Clinicians should actively involve family members in the care process, as these support networks can play a pivotal role in helping geriatric patients adhere to treatment plans and self-care regimens (31). Tailored support groups designed to address the unique emotional and practical needs of geriatric individuals with diabetes can provide an additional layer of assistance and guidance. Furthermore, establishing a strong physician-patient relationship characterized by trust and collaboration is essential (30). Clinicians should prioritize effective communication, active listening, and shared decision-making. By understanding and respecting the values and preferences of geriatric patients, healthcare providers can tailor treatment plans that align with individual needs, ultimately enhancing patient confidence and cooperation.

In the psychological domain, individualized support is important, cognitive-behavioral strategies, regular psychological check-ins, and coping skills education to manage emotional aspects of diabetes, including anxiety, depression, and emotional adjustment (37). Social aspects are also key, focusing on reducing feelings of exclusion and fear of judgment through normalization and peer support groups (38). In the healthcare context, it is key to emphasize the essential qualities of reassurance and containment, stressing the need for building empathetic patient relationships, providing clear and accurate information, and addressing unique anxieties to enhance emotional well-being (39).

## Conclusion

This systematic review and meta-synthesis bring forth the voices of wisdom, presenting an insightful understanding of geriatric self-management of T2D in America. The themes that emerge from this synthesis underscore the significance of cultivating positive attitudes, acknowledging cultural diversity, and respecting individual values in healthcare practices. These voices, echoing the resilience and unique perspectives of geriatric individuals, underscore the significance of cultivating positive attitudes and acknowledging cultural diversity in healthcare practices. By addressing the barriers, respecting individual values, and embracing the wisdom of geriatric patients, we can enhance the self-management experience and, ultimately, improve the quality of life for the geriatric population living with T2D. This paper serves as a call for healthcare professionals and policymakers to adapt and tailor their approaches to meet the unique needs of the geriatric in their journey toward self-management of chronic diseases.

## Data Availability

All data produced in the present work are contained in the manuscript

## Acknowledgments

None

## References

1. Centers for Disease Control and Prevention. Diabetes Basics. Centers for Disease Control and Prevention. Available at: https://www.cdc.gov/diabetes/basics/diabetes.html#:~:text=About%2038%20million%20US%20adults,limb%20amputations%2C%20and%20adult%20blindness. Updated: September 5, 2023. Accessed November 20, 2023.

2. Lin J, Thompson TJ, Cheng YJ, et al. Projection of the future diabetes burden in the United States through 2060. Popul Health Metr. 2018;16(1):9. Published 2018 Jun 15. doi:10.1186/s12963-018-0166-4

3. Mather M, Scommegna P, Kilduff L. Fact Sheet: Aging in the United States. Population Reference Bureau. Available at: https://www.prb.org/resources/fact-sheet-aging-in-the-united-states/. Published July 15, 2019. Accessed November 20, 2023.

4. Keeler, L. W., & Bernstein, M. J. The future of aging in smart environments: Four scenarios of the United States in 2050. Futures. 2021;(133):102830. 10.1016/j.futures.2021.102830

5. Nations, United. “Growing at a Slower Pace, World Population Is Expected to Reach 9.7 Billion in 2050 and Could Peak at Nearly 11 Billion around 2100 | UN Desa Department of Economic and Social Affairs.” United Nations, United Nations, 17 June 2019, www.un.org/development/desa/en/news/population/world-population-prospects-2019.html.

6. Lunenfeld B, Stratton P. The clinical consequences of an ageing world and preventive strategies. Best Pract Res Clin Obstet Gynaecol. 2013;27(5):643–659. doi:10.1016/j.bpobgyn.2013.02.005

7. Sinclair AJ, Abdelhafiz AH. Challenges and Strategies for Diabetes Management in Community-Living Older Adults. Diabetes Spectr. 2020;33(3):217–227. doi:10.2337/ds20-0013

8. Goyal R, Singhal M, Jialal I. Type 2 Diabetes. [Updated 2023 Jun 23]. In: StatPearls [Internet]. Treasure Island (FL): StatPearls Publishing; 2023 Jan-. Available from: https://www.ncbi.nlm.nih.gov/books/NBK513253/

9. Nuha A. ElSayed, Grazia Aleppo, Vanita R. Aroda, Raveendhara R. Bannuru, et al.; on behalf of the American Diabetes Association, 13. Older Adults: Standards of Care in Diabetes—2023. Diabetes Care 1 January 2023; 46 (Supplement_1): S216–S229. 10.2337/dc23-S013

10. Corriere M, Rooparinesingh N, Kalyani RR. Epidemiology of diabetes and diabetes complications in the elderly: an emerging public health burden. Curr Diab Rep. 2013;13(6):805–813. doi:10.1007/s11892-013-0425-5

11. Cuddapah GV, Vallivedu Chennakesavulu P, Pentapurthy P, et al. Complications in Diabetes Mellitus: Social Determinants and Trends. Cureus. 2022;14(4):e24415. Published 2022 Apr 23. doi:10.7759/cureus.24415

12. Adu MD, Malabu UH, Malau-Aduli AEO, Malau-Aduli BS. Enablers and barriers to effective diabetes self-management: A multi-national investigation. PLoS One. 2019;14(6):e0217771. Published 2019 Jun 5. doi:10.1371/journal.pone.0217771

13. Hernandez-Tejada MA, Campbell JA, Walker RJ, Smalls BL, Davis KS, Egede LE. Diabetes empowerment, medication adherence and self-care behaviors in adults with type 2 diabetes. Diabetes Technol Ther. 2012;14(7):630–634. doi:10.1089/dia.2011.0287

14. Young BA, Lin E, Von Korff M, et al. Diabetes complications severity index and risk of mortality, hospitalization, and healthcare utilization. Am J Manag Care. 2008;14(1):15–23.

15. Saraf AA, Petersen AW, Simmons SF, et al. Medications associated with geriatric syndromes and their prevalence in older hospitalized adults discharged to skilled nursing facilities. J Hosp Med. 2016;11(10):694–700. doi:10.1002/jhm.2614

16. Chlebowy DO, Garvin BJ. Social support, self-efficacy, and outcome expectations: impact on self-care behaviors and glycemic control in Caucasian and African American adults with type 2 diabetes. Diabetes Educ. 2006;32(5):777–786. doi:10.1177/0145721706291760

17. Leung E, Wongrakpanich S, Munshi MN. Diabetes Management in the Elderly. Diabetes Spectr. 2018;31(3):245–253. doi:10.2337/ds18-0033

18. Chentli F, Azzoug S, Mahgoun S. Diabetes mellitus in elderly. Indian J Endocrinol Metab. 2015;19(6):744–752. doi:10.4103/2230-8210.167553

19. Chester B, Stanely WG, Geetha T. Quick guide to type 2 diabetes self-management education: creating an interdisciplinary diabetes management team. Diabetes Metab Syndr Obes. 2018;11:641–645. Published 2018 Oct 17. doi:10.2147/DMSO.S178556

20. Habebo TT, Pooyan EJ, Mosadeghrad AM, Babore GO, Dessu BK. Prevalence of Poor Diabetes Self-Management Behaviors among Ethiopian Diabetes Mellitus Patients: A Systematic Review and Meta-Analysis. Ethiop J Health Sci. 2020;30(4):623–638. doi:10.4314/ejhs.v30i4.18

21. Okoye OC, Ohenhen OA. Assessment of diabetes self-management amongst Nigerians using the diabetes self-management questionnaire: a cross-sectional study. Pan Afr Med J. 2021;40:178. Published 2021 Nov 24. doi:10.11604/pamj.2021.40.178.28584

22. BMJ (OPEN ACCESS) Page MJ, McKenzie JE, Bossuyt PM, Boutron I, Hoffmann TC, Mulrow CD, et al. The PRISMA 2020 statement: an updated guideline for reporting systematic reviews. BMJ 2021;372:n71. doi: 10.1136/bmj.n71

23. Tong, A., Flemming, K., McInnes, E. et al. Enhancing transparency in reporting the synthesis of qualitative research: ENTREQ. BMC Med Res Methodol 2012;12 (181). 10.1186/1471-2288-12-181

24. CASP UK. CASP systematic review checklist. Oxford: CASP UK. 2018. Available from: https://casp-uk.net/wp-content/uploads/2018/01/CASP-Systematic-Review-Checklist_2018.pdf

25. Butler A, Hall H, Copnell B. A Guide to Writing a Qualitative Systematic Review Protocol to Enhance Evidence-Based Practice in Nursing and Health Care. Worldviews Evid Based Nurs. 2016;13:241–249

26. Washington G, Wang-Letzkus MF. Self-care practices, health beliefs, and attitudes of older diabetic Chinese Americans. J Health Hum Serv Adm. 2009;32(3):305–323.

27. Chlebowy DO, Hood S, LaJoie AS. Facilitators and barriers to self-management of type 2 diabetes among urban African American adults: focus group findings. Diabetes Educ. 2010;36(6):897–905. doi:10.1177/0145721710385579

28. George, S.R. and Thomas, S.P. (2010), Lived experience of diabetes among older, rural people. Journal of Advanced Nursing, 66: 1092–1100. 10.1111/j.1365-2648.2010.05278.x

29. Kirk A, Mutrie N, MacIntyre P, Fisher M. Increasing physical activity in people with type 2 diabetes. Diabetes Care. 2003;26(4):1186–1192. doi:10.2337/diacare.26.4.1186

30. Beverly EA, Fitzgerald S, Sitnikov L, Ganda OP, Caballero AE, Weinger K. Do older adults aged 60–75 years benefit from diabetes behavioral interventions? Diabetes Care. 2013;36(6):1501–1506. doi:10.2337/dc12-2110.

31. Joo JY, Lee H. Barriers to and facilitators of diabetes self-management with elderly Korean-American immigrants. Int Nurs Rev. 2016;63(2):277–284. doi:10.1111/inr.12260

32. Bustillos BD, Sharkey JR. "I Try to Keep That Sugar Down." Experiences of Homebound Older Adults With Type 2 Diabetes: Barriers to Self-Management. J Nutr Gerontol Geriatr. 2020;39(1):69-87. doi:10.1080/21551197.2019.1695037

33. Large S. Type 2 Diabetes Mellitus, Cognition And Self-Management Behavior In Mexican American Older Adults. Graduate School of Texas Women’s University. 2022.

34. Wilson, C, Stock, J. The impact of living with long-term conditions in young adulthood on mental health and identity: What can help? Health Expect. 2019; 22: 1111–1121. 10.1111/hex.12944

35. Dayer L, Heldenbrand S, Anderson P, Gubbins PO, Martin BC. Smartphone medication adherence apps: potential benefits to patients and providers. J Am Pharm Assoc (2003). 2013;53(2):172-181. doi:10.1331/JAPhA.2013.12202

36. Herges JR, Neumiller JJ, McCoy RG. Easing the Financial Burden of Diabetes Management: A Guide for Patients and Primary Care Clinicians. Clin Diabetes. 2021;39(4):427–436. doi:10.2337/cd21-0004

37. Kalra S, Jena BN, Yeravdekar R. Emotional and Psychological Needs of People with Diabetes. Indian J Endocrinol Metab. 2018;22(5):696–704. doi:10.4103/ijem.IJEM_579_17

38. Stuart H. Reducing the stigma of mental illness. Glob Ment Health (Camb). 2016;3:e17. Published 2016 May 10. doi:10.1017/gmh.2016.11

39. Moudatsou M, Stavropoulou A, Philalithis A, Koukouli S. The Role of Empathy in Health and Social Care Professionals. Healthcare (Basel). 2020;8(1):26. Published 2020 Jan 30. doi:10.3390/healthcare8010026

40. Li TJ, Zhou J, Ma JJ, Luo HY, Ye XM. What are the self-management experiences of the elderly with diabetes? A systematic review of qualitative research. World J Clin Cases. 2022 Feb 6;10(4):1226–1241.

41. CASP. (2018). CASP-Critical Appraisal Skills Programme. CASP-Critical Appraisal Skills Programme website. Retrieved from https://casp-uk.net/

42. Narindrarangkura P, Boren SA, Khan U, Day M, Simoes EJ, Kim MS. SEE-diabetes, a patient-centered diabetes self-management education and support for older adults: Findings and information needs from providers’ perspectives. Diabetes Metab Syndr. 2022;16(9):102582. doi:10.1016/j.dsx.2022.102582

43. U.S. Census Bureau. Racial and Ethnic Diversity in the United States: 2010 and 2020 Census. Published August 12, 2021 [cited 2024 April 4]. Available from: https://www.census.gov/library/visualizations/interactive/racial-and-ethnic-diversity-in-the-united-states-2010-and-2020-census.html

44. Vespa J, Medina L, Armstrong DM. Demographic Turning Points for the United States: Population Projections for 2020 to 2060. Current Population Reports. P25–1144. U.S. Census Bureau, Washington, DC; 2020. Available from: https://www.census.gov/content/dam/Census/library/publications/2020/demo/p25-1144.pdf

45. U.S. Census Bureau. Census Bureau Reports Majority of Births Occurring Outside of Marriage for Select Groups. Published December 6, 2012 [cited 2024 April 4]. Available from: https://www.census.gov/newsroom/releases/archives/population/cb12-243.html#:~:text=Of%20those%20age%2065%20and,12.5%20percent%20non%2DHispanic%20black

46. Pandya N, Hames E, Sandhu S. Challenges and Strategies for Managing Diabetes in the Elderly in Long-Term Care Settings. Diabetes Spectr. 2020;33(3):236–245. doi:10.2337/ds20-0018

